# The Morbidity Costs of Air Pollution through the Lens of Health Spending in China

**DOI:** 10.1101/2021.09.11.21256274

**Authors:** Xin Zhang, Xun Zhang, Xi Chen, Yuehua Liu, Xintong Zhao

## Abstract

This study offers one of the first causal evidence on the morbidity costs of fine particulates (PM2.5) for all age cohorts in a developing country, using individual-level healthcare spending data from the basic medical insurance program in Wuhan, China. Our instrumental variable (IV) approach uses thermal inversion to address potential endogeneity in PM2.5 concentrations and shows that PM2.5 imposes a significant impact on medical expenditures. The IV estimate suggests that a 10 μg/m^3^ reduction in monthly average PM2.5 leads to a 2.79% decrease in the value of health spending and a 0.70% decline in the number of transactions in pharmacies and health facilities. The effect is more salient for males, children, and older adults. Moreover, our estimates provide a lower bound of people’s willingness-to-pay, which amounts to CNY 51.85 (or USD 8.38) per capita per year for a 10 μg/m^3^ reduction in PM2.5.

**JEL Codes:** Q51, Q53, I11, I31

**Conflict of interest:** The authors declare that they have no conflict of interest.

## 1. Introduction

A large body of literature examines the effects of air pollution on health-related outcomes, including adult mortality (Anderson 2019; Barreca, Neidell, and Sanders 2017; Chay, Dobkin, and Greenstone 2003; He, Fan, and Zhou 2016); infant mortality (Chay and Greenstone 2003; Greenstone and Hanna 2014; Knittel, Miller, and Sanders 2016; Luechinger 2014; Tanaka 2015); hospitalization (Moretti and Neidell 2011; Schlenker and Walker 2016); life expectancy (Chen et al. 2013; Ebenstein et al. 2017); and birth outcomes (Currie and Neidell 2005; Currie, Neidell, and Schmieder 2009). However, evidence on the causal effect of air pollution on medical expenditures is relatively limited. The morbidity costs of air pollution provide a lower bound estimate on individuals’ willingness–to–pay (WTP) for better air quality, a requisite for the government to conduct cost–benefit analyses when introducing more optimal environmental regulations.

While there has been a large set of epidemiological studies on the association between air pollution and health spending, it remains important to carefully design causal evaluations in examining the effect of air pollution on medical expenditures in order to address sources of bias due to the endogeneity problem. The first source is unobserved factors. Time-varying local shocks may be correlated with both health spending and exposure to air pollution, and they cannot be fully removed by individual fixed effects and time-fixed effects. The second source relates to avoidance behavior or residential sorting. In recent years, air pollution has attracted greater public awareness in China. On days when pollution levels are high, residents may reduce outdoor activities (Neidell 2009), postpone visits to health care facilities, or take preventive measures by wearing particulate-filtering facemasks or using air-filtering products (Ito and Zhang 2019; Sun, Kahn, and Zheng 2017; Zhang and Mu 2018). In the long term, those more vulnerable to air pollution may even migrate to avoid more polluted areas (Chen, Oliva, and Zhang 2017). The third source, i.e., measurement errors with regard to air pollutants, may be attributable to aggregation of pollution data from sporadic monitoring stations or data manipulation (Chen et al. 2012; Ghanem and Zhang 2014).

Only a few studies have attempted to identify the causal effect employing plausibly exogenous variations. Deschênes, Greenstone, and Shapiro (2017) examine the effect of a decline in nitrogen oxides (NO_X_) emissions on pharmaceutical expenditures at the county-season-year level, employing a cap-and-trade NO_X_ Budget Program (NBP) as a quasi-experiment. Instrumenting air pollution using changes in local wind directions, Deryugina et al. (2019) estimate the causal effects of daily fine particulates (PM2.5) on county-level inpatient emergency room (ER) spending for those aged 65 years and older. Williams and Phaneuf (2019) identify the impact of PM2.5 on quarterly household health spending via instrumenting local air pollution using emissions from distant sources. Utilizing a similar instrumental variable (IV) strategy, Barwick et al. (2018) first analyze the medical burden from PM2.5 in a developing country, based on city-level credit and debit card transactions in China. Their IV estimates suggest that a 10 μg/m^3^ increase in PM2.5 leads to a 2.65% increase in the number of transactions and a 1.5% increase in the value of transactions. Liu and Ao (2021) estimate the impact of daily air quality index (AQI) on outpatient health care expenditures for respiratory diseases at the township level using thermal inversion as an IV. Employing a similar IV, Liao, Du, and Chen (2021) study the effect of PM2.5 on people’s yearly medical expenses obtained from a social survey in China.

In this paper, we examine the causal impact of PM2.5 exposure on medical expenses and estimate people’s WTP for cleaner air, matching individual-level health spending data with air pollution exposure between 2013 and 2015. We instrument PM2.5 concentrations using thermal inversion, a widely-used IV in recent studies (Arceo, Hanna, and Oliva 2016; Chen, Guo, and Huang 2018; Chen et al. 2017; Liao, Du, and Chen 2021) to address potential endogeneity in air pollution. Thermal inversion is a meteorological phenomenon that occurs when air temperature is abnormally higher than that at lower altitudes. It reduces vertical circulation of the air and thus traps air pollutants near the ground. While thermal inversion does not pose a direct threat to health, it does lead to higher concentrations of air pollution (Arceo et al. 2016). Our results suggest that a 10 μg/m^3^ reduction in monthly average PM2.5 leads to a 2.79% decrease in the value of health spending, and a 0.70% decrease in the number of transactions in pharmacies and health facilities. This effect is more salient for males, children (up to age 5), and older people (age 51 and older) who are in poor health. The strong effect on pharmaceutical spending highlights the importance to incorporate in analyses data on health spending from both pharmacies and all levels of health facilities. Valuing air quality using total health spending data, our estimates suggest that people are willing to pay 51.85 Chinese *yuan* (CNY) per capita per year for a 10 μg/m^3^ reduction in PM2.5.

We contribute to the literature in several dimensions. First, we use individual-level data to identify medical spending in response to air pollution. Previous studies often rely on aggregated health spending data that are subject to the *ecological fallacy*. The findings may be biased, depending on the level of aggregation, due to the omitted variables that often threaten identification in research linking air pollution to behavior outcomes. Employing comprehensive individual-level data also enables us to test the heterogeneous effects across groups to understand which segments of population are more affected by air pollution. To our best knowledge, only two economics studies examine the effects of air pollution on medical expenditures at the disaggregated level (Liao, Du and Chen 2021; Williams and Phaneuf 2019). However, they all obtain healthcare spending and utilization data from social surveys, which may suffer from recall error or other potential biases.

Second, some time–invariant unobserved factors—such as individuals’ health stock and preference to live in a clean environment—are correlated with both health spending and exposure to air pollution, thereby biasing the estimations. By exploiting the longitudinal nature of our health spending data at the individual level, we are among the first to control for individual fixed effects in our estimation of the morbidity costs of air pollution, thereby mitigating concerns over individual heterogeneity in preference or other unobservables (Deschênes et al. 2017).

Third, existing studies either focus on middle-aged or older adults or make no distinction by patient age. Since our sample covers all age cohorts of urban residents with precise information on patient age, we could perform more generalized heterogeneity analysis across all age groups to understand what age groups are more sensitive to air pollution. Specifically, we examine the impact of air pollution on medical spending of younger individuals, a group less studied in the literature.

Fourth, most studies either use inpatient medical expenditures (Liao et al. 2021) or total inpatient and outpatient expenses (Barwick et al. 2018; Deschênes et al. 2017). However, inpatient health transactions are likely scheduled in advance and thus immune to transitory air pollution. For example, Barwick et al. (2018) find no significant effects of air pollution on total health spending due to the large noise inherent in the value of total medical expenditures. They instead employ the number of health-related transactions in estimating the morbidity cost of PM2.5. Fortunately, as we possess detailed information on each category of transactions in pharmacies and all levels of health facilities, we focus on outpatient related expenses data, which have been less examined in economics studies.

Moreover, our paper contributes to the strand of literature that estimates individuals’ WTP for improved air quality. ^1^ Following the health production framework established in the seminal work Grossman (1972), Deschênes et al. (2017), Williams and Phaneuf (2019) and Barwick et al. (2018) propose a theoretical model of WTP and show that the benefits accrued to reduced health spending is merely one component of people’s WTP for improved air quality. Therefore, our estimated WTP using medical expenditures may offer a lower bound of WTP for cleaner air.^2^

Finally, we are among the first to estimate the morbidity costs of air pollution in a developing country using health spending records for all age cohorts. Air pollution is generally worse in developing countries, such as Nepal, Bangladesh, India, China, and Pakistan.^3^ In fact, 98.6% of the total population in China were exposed to PM2.5 at unsafe levels according to the World Health Organization (WHO) guideline (Long et al. 2018). We obtain health spending data from Wuhan, the capital city of Hubei province, China. As a major manufacturing city in central China, Wuhan is exposed to high levels of air pollution with large daily variations. Therefore, the dose–response relationship between air pollution and medical expenses estimated in this study may have implications for other developing countries.

The remainder of this paper is organized as follows. Section 2 describes data sources. Section 3 discusses the empirical model and the identification strategy that uses thermal inversion as an instrument. Section 4 reports the main findings, including baseline results, robustness checks, and heterogeneous effects. Section 5 compares our calculated WTP to others in the related literature. Section 6 concludes and discusses future research directions.

## 2. Data

### 2.1. Health spending

Health spending data are obtained from the universal basic medical insurance system, developed by the Chinese central government and covering 95% of Chinese population as of 2011 (Yu 2015). The system includes two government programs in urban areas—namely, Urban Employee Basic Insurance (UEBMI) and Urban Resident Basic Medical Insurance (URBMI).^4^ We use a 1% representative sample from the basic medical insurance program in urban areas of Wuhan, the capital city of Hubei province, China. Our dataset includes all the health expenditure records for 120 health facilities (i.e., clinics and hospitals), more than 3,000 pharmacies, and approximately 40,000 patients between 2013 and 2015. ^5^ For each record, we observe patient unique ID, gender, age, location of health facilities, time, and total value of expenses. Total health spending includes outpatient and inpatient expenses at both pharmacies and all levels of health facilities. Most inpatient health transactions are likely related to surgeries, with appointments made in advance, and thus are insensitive to transitory air pollution. Therefore, we utilize only outpatient health spending in our analysis. Pharmacies and health facilities account for 39.6% and 60.4% of total health spending in our data, respectively. Medical expenses are further classified into three categories: medication, examination, and treatment. Figure A1 plots both the weekly values of health spending and the number of transactions from 2013 to 2015. Medical spending and number of transactions decline during holidays, especially the Spring Festivals.

For our purposes, there are four advantages in employing data from the basic medical insurance program in Wuhan. First, the program provides wide coverage in Wuhan, and the beneficiaries in our sample cover all age groups of urban residents, enabling us to examine heterogeneous effects across age cohorts and to understand which subpopulations are most affected by air pollution. Second, the health spending records in our sample include all pharmacy and health facility transactions, enabling us to estimate the morbidity costs of air pollution in a more comprehensive way than can studies that consider only medical expenses incurred in hospitals. Third, information on geographic locations of pharmacies and health facilities, as well as dates of service, enable us to precisely match individual-level healthcare expenditures with external air quality data. Fourth, the daily mean concentration of PM2.5 in Wuhan in 2013–2015 was 80 μg/m^3^, a much higher figure than that in developed countries. This useful setting provides us an opportunity not only to examine the non-linear effect of PM2.5 on health spending, but also to estimate a wide range of dose–response relationships.

### 2.2. Pollution and weather

Air pollution measures are provided by the daily air quality report of the Ministry of Ecology and Environment (MEE) of China, which started to publish concentrations of six air pollutants and an air quality index (AQI) in 2013.^6^ It covers 10 monitoring stations in Wuhan city, with the longitudes and latitudes of each station provided. Given that PM2.5 are more toxic and can penetrate deeper into lungs than PM10, we mainly focus on PM2.5 (Pope and Dockery 2006).^7^ Figure A2 shows the daily mean PM2.5 in Wuhan during the period from 2013 to 2015. From Figure A2, on most days, the concentrations of PM2.5 are higher than the daily air quality guideline values of the WHO (25 μg/m^3^).

The weather data originates from the China National Meteorological Data Service Center (CMDC), part of the National Meteorological Information Center of China. The dataset reports consecutive daily records on a wide range of weather conditions, including temperature, precipitation, wind speed, sunshine duration, and relative humidity. We calculate the mean values for each weather variable from two monitoring stations in Wuhan.

### 2.3. Thermal inversion

The data on thermal inversion is drawn from the product M2I6NPANA, version 5.12.4, released by the U.S. National Aeronautics and Space Administration (NASA). The data reports air temperatures every six hours for each 0.5° × 0.625° (around 50 km × 60 km) grid, for 42 layers, ranging from 110 meters to 36,000 meters. We aggregate all the grids within Wuhan and calculate their mean values. For every six-hour period, we further derive the temperature difference between the second layer (320 meters) and the first layer (110 meters). Under normal conditions, the difference would be negative, since temperature decreases as latitude increases. However, thermal inversion occurs when the temperature difference is positive. If the difference is positive, the magnitude measures the thermal inversion strength. If the difference is negative, we truncate it to zero. Following Chen, Guo, and Huang (2018), we define a daily thermal inversion if the temperature difference is positive for any six-hour period during the 24 hours from 6 pm on the previous day until 6 pm on the current day, and average the thermal inversion strength across the four six-hour periods for each day accordingly. Then we calculate the mean value of thermal inversion strength and the probability of occurrence within each month based on the daily data.

In order to match air pollution measures with the health spending data, we infer the home address for each person from the location of the pharmacy that one most often visited, and match the monthly mean concentrations from the nearest monitoring station. Figure 1 plots the distribution of monitoring stations and health facilities in Wuhan. For weather and thermal inversion data, we match their monthly mean values to each individual.

**Figure 1:**
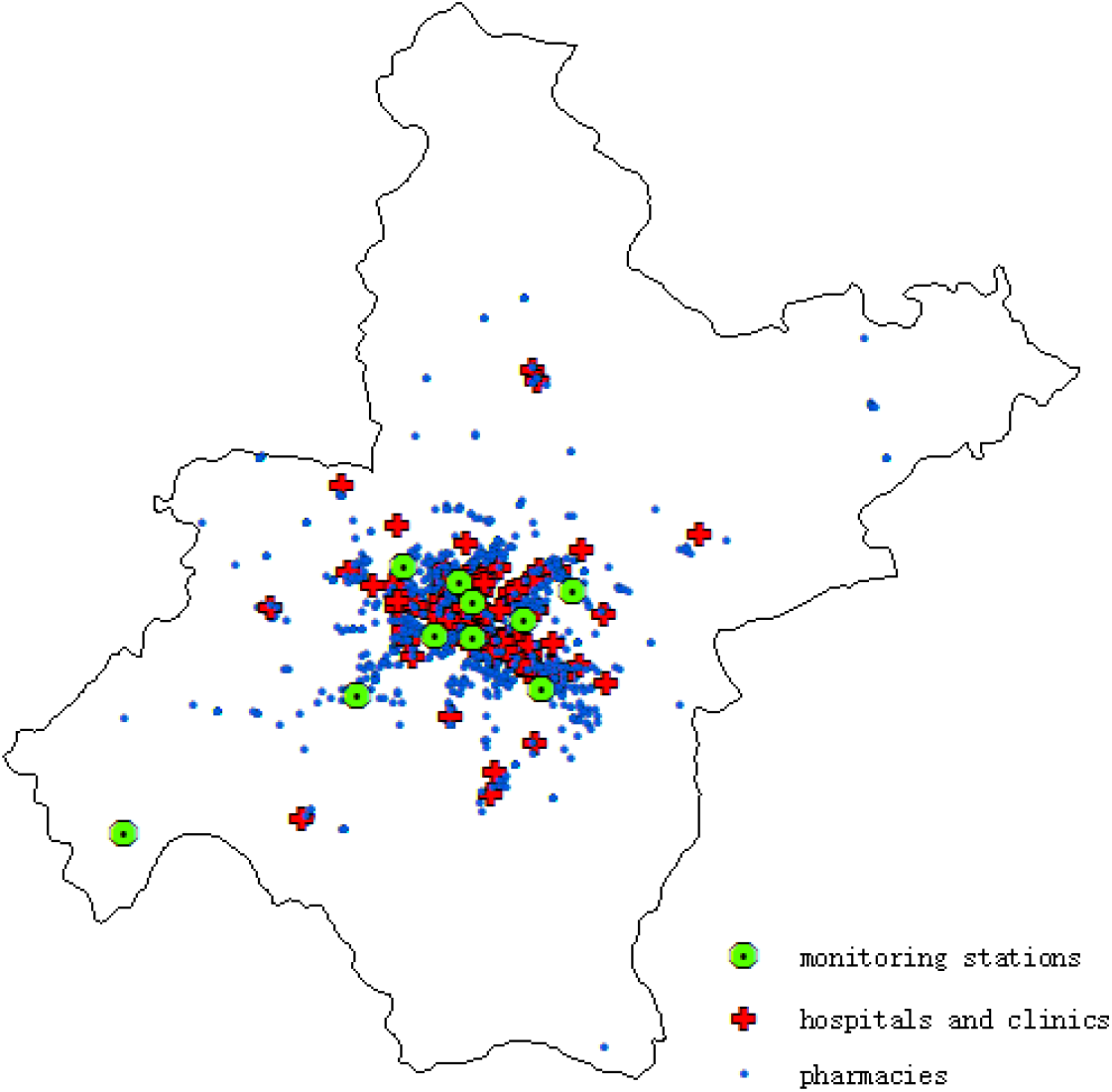
Distribution of monitoring stations and health facilities in Wuhan, China.

Our dataset contains around 1.38 million transactions in pharmacies and all levels of health facilities from 40,000 individuals. We calculate the total value of health spending as well as the number of transactions in pharmacies and health facilities for each person in each month. If no health expenditure records exist for one specific month, we assign a value of zero. In this way, we construct a data panel at the individual-month level. The final dataset for analysis includes over 1.45 million person-month observations.

## 3. Empirical strategy

Our baseline econometric specification is:

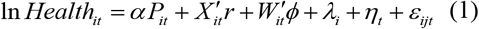

The dependent variable *Health*_*it*_ is the value of health spending, or the number of transactions in pharmacies and health facilities for individual *i* during month *t*. The key variable *P*_*it*_ represents the mean concentration of PM2.5 for individual *i* during month *t*. We include a set of demographic controls *X*_*it*_, including age and its squared term. We also control for a vector of rich weather conditions *W*_*t*_, involving temperature bins (<45°F, 45–55°F, 55–65°F, 65–75°F, and >75°F), precipitation, wind speed, sunshine duration, and relative humidity in square polynomial forms to mitigate the concern that they are correlated with both health spending and air quality. *λ*_*i*_ represents individual fixed effects, while *η*_*t*_ indicates year and month fixed effects. *ε*_*ijt*_ is the error term. To address both spatial and serial correlation over time, standard errors are clustered at the county-year-month level.^8^ Table 1 displays key variables and their summary statistics.

**Table 1:** Summary statistics of key variables.

| **Variable** | **Mean** | **Std. Dev.** |
| --- | --- | --- |
| **Insurance claim data** |  |  |
| value of health spending per month, *yuan* | 154.86 | 1026.00 |
| number of transactions per month | 0.937 | 1.576 |
| male | 0.492 | 0.500 |
| age | 42.168 | 19.051 |
| **Air pollution** |  |  |
| PM2.5 concentration, $\mu\text{g}/\text{m}^3$ | 80.790 | 40.639 |
| **Thermal inversion** |  |  |
| strength, $^{\circ}\text{C}$ | 0.279 | 0.184 |
| probability of occurrence, % | 0.546 | 0.178 |
| **Weather** |  |  |
| temperature in: | 0.250 | 0.433 |
| <45 $^{\circ}\text{F}$ | 0.111 | 0.315 |
| 45-55 $^{\circ}\text{F}$ | 0.139 | 0.346 |
| 55-65 $^{\circ}\text{F}$ (*reference group*) | 0.194 | 0.396 |
| 65-75 $^{\circ}\text{F}$ | 0.306 | 0.461 |
| >75 $^{\circ}\text{F}$ | 3.744 | 2.607 |
| precipitation, *cm* | 2.014 | 0.289 |
| wind speed, *m/s* | 4.919 | 1.698 |
| sunshine duration, *hour* | 75.652 | 5.816 |
| relative humidity, % | 154.86 | 1026.00 |

OLS estimates of equation (1) are prone to bias resulting from potential sources of endogeneity, such as time-varying unobserved factors, avoidance behaviors, and measurement error in air pollution due to the aggregation of pollution data from sporadic monitoring stations at the city level.^9^ We address endogeneity by employing an IV strategy, using thermal inversion as an instrument for air pollution. Thermal inversion is a common phenomenon that occurs when a layer of hot air covers a layer of cooler air near the ground. It prevents air flow by trapping air pollutants in the lower atmosphere and has no adverse effects on human health (Arceo et al. 2016). We take advantage of this exogenous shock in order to identify the effects of air pollution on health spending.

The specification for our first stage is:

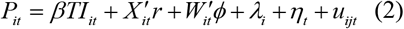

The excluded instrument is *TI*_*it*_, which is the average thermal inversion strength for individual *i* during month *t*. Thermal inversion strength is defined as the air temperature at the second layer (320 meters) minus the temperature near the ground (110 meters). We keep the positive differences and truncate the negative differences to zero. Standard errors are clustered at the county-year-month level. Other control variables are as defined in equation (1).

Figure 2 illustrates the relationships in Wuhan between thermal inversion strength and PM2.5 at the monthly level during the sample period. As shown by the figures, thermal inversion strength is highly correlated with PM2.5 concentrations, indicating that thermal inversion is a strong predictor of air pollution levels.

**Figure 2:**
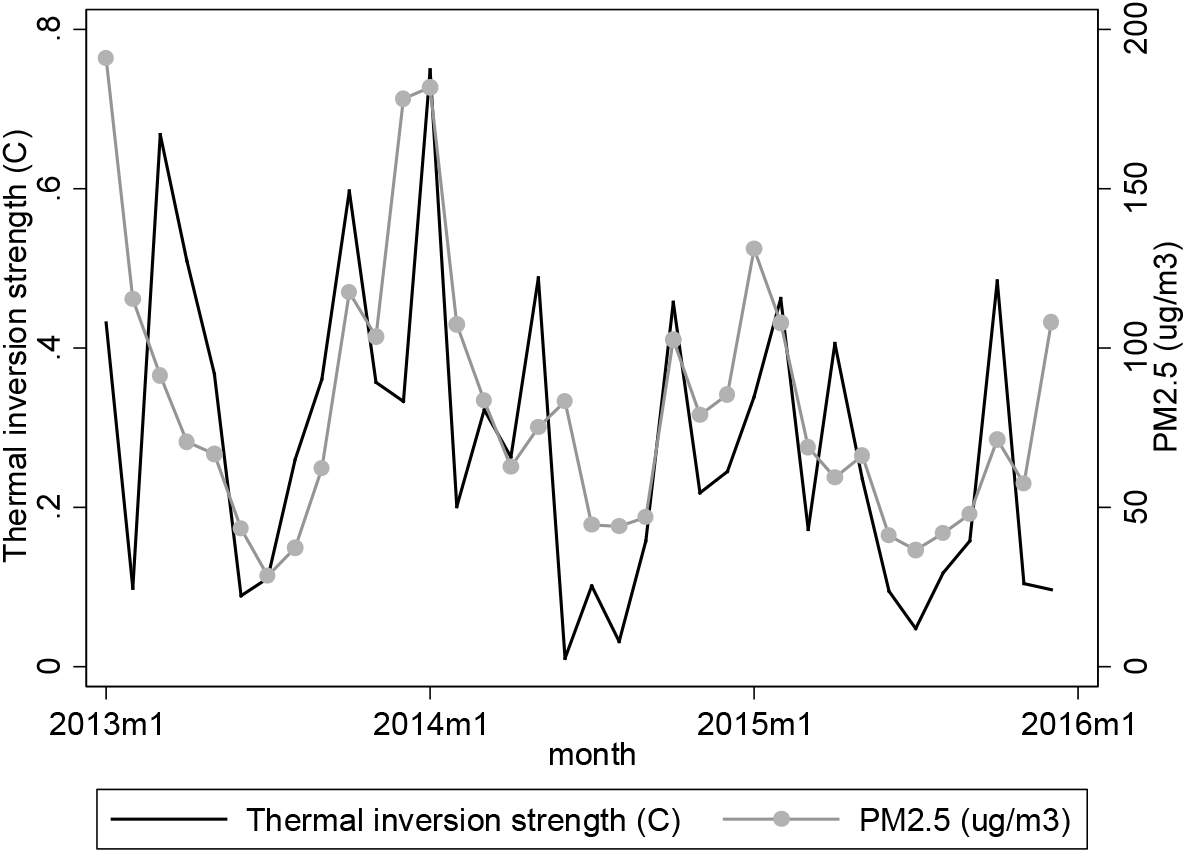
Monthly trend of thermal inversion strength and PM2.5 in Wuhan, 2013-2015.

Our second stage model is specified as:

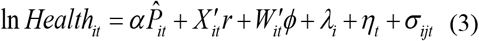

The variable 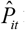 is the predicted value for *P*_*it*_from the first stage. Standard errors are clustered at the county-year-month level. The other control variables are defined as in equation (1). After flexibly controlling for a large number of fixed effects and covariates, our identification assumption is that changes in a city’s thermal inversion are unrelated to changes in health spending, except through air pollution.

Before undertaking quantitative analyses, we plot the relationship between PM2.5 and health spending outcomes. As shown in Figure 3, the value of health spending, as well as the number of transactions in pharmacies and health facilities, is slightly positively correlated with PM2.5 levels. Of course, these bivariate plots provide suggestive evidence only. More rigorous analyses are needed to control for other confounding factors.

**Figure 3:**
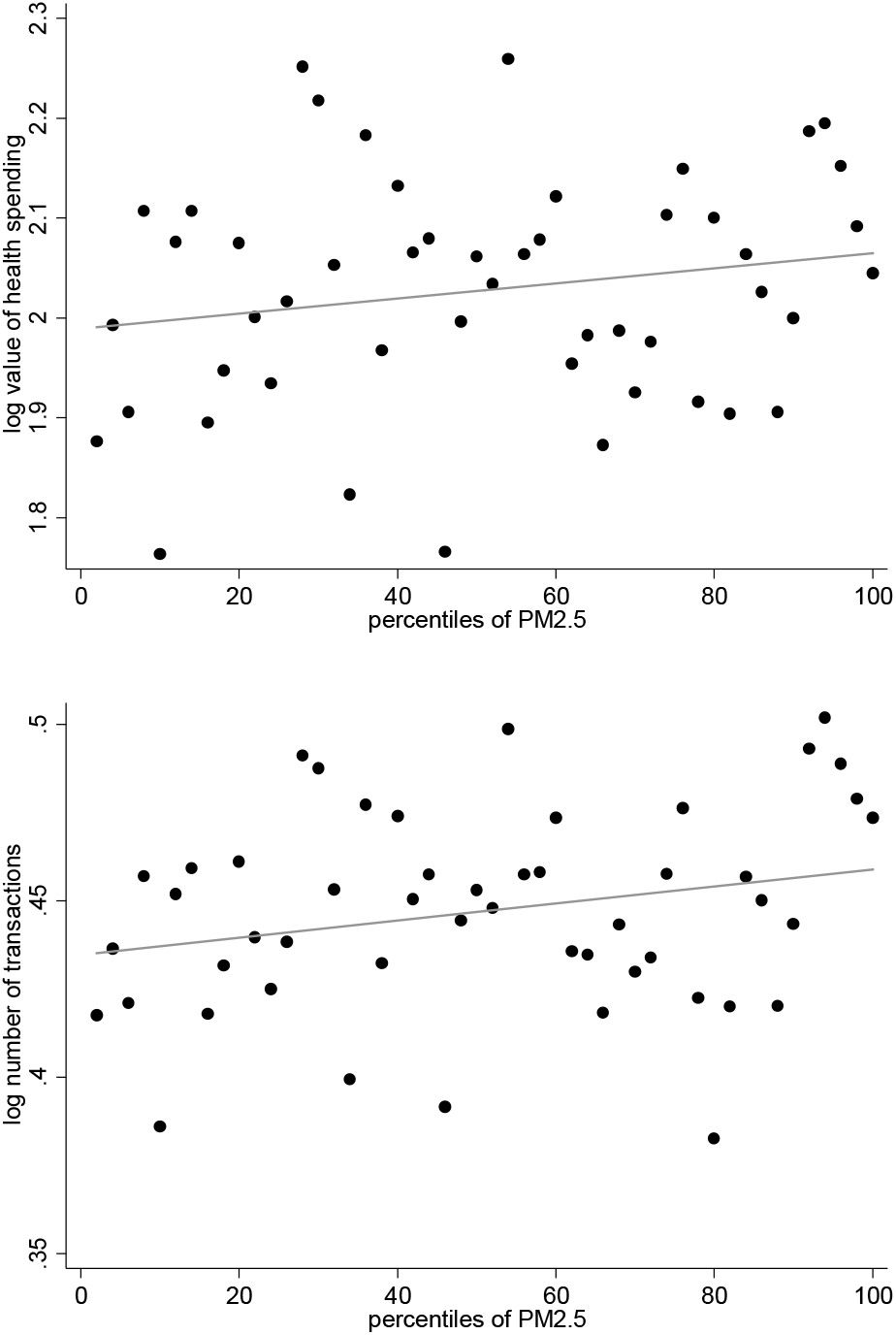
Relationship between health spending outcomes and PM2.5 concentrations. Note: Each dot denotes the in-group average of the health spending outcomes. Groups are binned by percentiles of the x-axis variable, PM2.5.

## 4. Results

### 4.1. Baseline results

Table 2 presents the first-stage estimates of the effect of thermal inversions on PM2.5 concentrations. The regression controls for individual fixed effects, demographic controls, weather controls, as well as year and month fixed effects. As the magnitude of the coefficient is difficult to interpret, we convert the point estimate to elasticity. The point estimate indicates that a 1 percent increase in thermal inversion is associated with a 0.16 percent (46.196 × 0.279/80.790) increase in PM2.5 concentrations. Overall, we find a strong first-stage relationship. The Kleibergen-Paap (KP) F-statistics is well above the Stock-Yogo critical value.

**Table 2:** Effects of thermal inversion on air pollution (first stage)

| Dependent variable | PM2.5, $\mu\text{g}/\text{m}^3$ |
| --- | --- |
| Thermal inversion strength, $^{\circ}\text{C}$ | 46.196***<br>(10.278) |
| demographic controls | Yes |
| weather controls | Yes |
| individual fixed effects | Yes |
| year and month fixed effects | Yes |
| KP first-stage F-statistic | 20.20 |
| Observations | 1,447,649 |

Note: The demographic controls include age and its square term. The weather controls include temperature bins (<45 $^{\circ}\text{F}$ , 45-55 $^{\circ}\text{F}$ , 55-65 $^{\circ}\text{F}$ , 65-75 $^{\circ}\text{F}$ , >75 $^{\circ}\text{F}$ ), total precipitation, mean wind speed, sunshine duration and relative humidity in polynomial forms. Robust standard errors, clustered at the county-year-month level, are presented in parentheses. \*10% significance level. \*\*5% significance level. \*\*\*1% significance level.

Table 3 shows our baseline results of the effects of air pollution on value of health spending in Panel A and number of transactions in Panel B. Columns (1) and (3) report the OLS estimates of equation (1), while columns (2) and (4) report the IV estimates. The OLS estimate in Column (1) suggests a significant positive correlation between PM2.5 and value of health spending after controlling individual fixed effects, demographic controls, weather controls, and year and month fixed effects. The point estimate indicates that a 1 μg/m^3^ reduction in monthly average PM2.5 leads to a decrease in medical expenditures by 0.754‰. As the marginal effect of exposure to air pollution on health spending provides a lower bound of people’s WTP for better air quality, the coefficient on PM2.5 indicates that people are, on average, willing to allocate 0.754% of their medical expenditure to a 10 μg/m^3^ monthly reduction in PM2.5. To put this into context, note that the mean of monthly health spending is CNY 154.86. The WTP amounts to CNY 14.01 per year (= 0.754% × 154.86 × 12) for a 10 μg/m^3^ reduction in PM2.5. These two numbers are reported in the last two rows of Table 3.

**Table 3:**
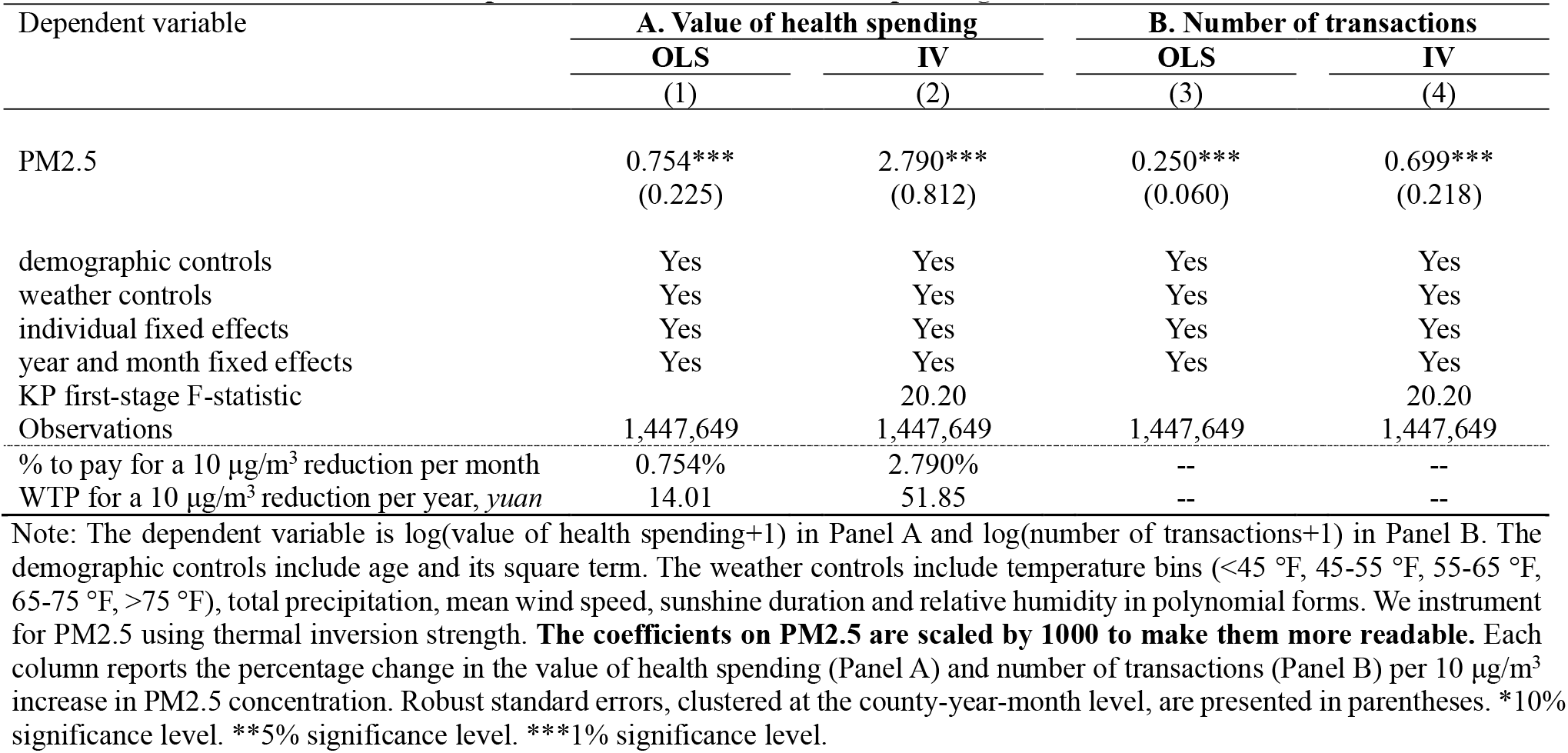
Effects of air pollution on the value of health spending and number of transactions.

| Dependent variable | A. Value of health spending |  | B. Number of transactions |  |
| --- | --- | --- | --- | --- |
|  | OLS | IV | OLS | IV |
|  | (1) | (2) | (3) | (4) |
| PM2.5 | 0.754***<br>(0.225) | 2.790***<br>(0.812) | 0.250***<br>(0.060) | 0.699***<br>(0.218) |
| demographic controls | Yes | Yes | Yes | Yes |
| weather controls | Yes | Yes | Yes | Yes |
| individual fixed effects | Yes | Yes | Yes | Yes |
| year and month fixed effects | Yes | Yes | Yes | Yes |
| KP first-stage F-statistic |  | 20.20 |  | 20.20 |
| Observations | 1,447,649 | 1,447,649 | 1,447,649 | 1,447,649 |
| % to pay for a 10 $\mu\text{g}/\text{m}^3$ reduction per month | 0.754% | 2.790% | -- | -- |
| WTP for a 10 $\mu\text{g}/\text{m}^3$ reduction per year, *yuan* | 14.01 | 51.85 | -- | -- |

Note: The dependent variable is $\log(\text{value of health spending}+1)$ in Panel A and $\log(\text{number of transactions}+1)$ in Panel B. The demographic controls include age and its square term. The weather controls include temperature bins (<45 °F, 45-55 °F, 55-65 °F, 65-75 °F, >75 °F), total precipitation, mean wind speed, sunshine duration and relative humidity in polynomial forms. We instrument for PM2.5 using thermal inversion strength. **The coefficients on PM2.5 are scaled by 1000 to make them more readable.** Each column reports the percentage change in the value of health spending (Panel A) and number of transactions (Panel B) per 10 $\mu\text{g}/\text{m}^3$ increase in PM2.5 concentration. Robust standard errors, clustered at the county-year-month level, are presented in parentheses. \*10% significance level. \*\*5% significance level. \*\*\*1% significance level.

Columns (2) presents the corresponding IV estimate of the causal effect of PM2.5 on value of health spending. The IV estimate is approximately four times larger than the corresponding OLS estimate, suggesting that OLS estimation suffers from significant bias. Furthermore, the IV estimate implies that people are, on average, willing to pay CNY 51.85 per year (= 2.790% × 154.86 × 12) for a 10 μg/m^3^ reduction in PM2.5.

A large difference between the OLS and IV results on the effects of air pollution on health spending is common in the literature (Barwick et al. 2018; Deryugina et al. 2019; Williams and Phaneuf 2019). Two possible reasons exist for this downward bias. First, some time-varying omitted variables, such as economic growth and infrastructure construction, are positively correlated with air pollution. As these factors are likely to reduce health spending, the omitted-variable bias is negative. Second, measurement error in PM2.5 could lead to attenuation bias.

Panel B of Table 3 shows the effect of exposure to PM2.5 on the number of transactions at the monthly level. Similarly, the IV estimates are larger than the corresponding OLS estimates, indicating that OLS estimation still suffers from downward bias. As presented in Column (4), the IV estimate suggests that a 10 μg/m^3^ reduction in monthly average PM2.5 leads to a 0.70% decrease in the number of transactions in pharmacies and health facilities.

### 4.2. Robustness checks

In this section, we conduct a set of regressions to check the robustness of our main results. The first issue is that the concentrations of various air pollutants are highly correlated, and thus the estimation of WTP for PM2.5 reduction may involve payment to other co-pollutants. ^10^ In order to address this concern, we add co-pollutants, including PM2.5-10, CO, O_3_, SO_2,_ and NO_2_, respectively. As PM10 contains PM2.5, we add PM2.5-10, which represents particulate matter with a diameter between 2.5 to 10 μm, as a co-pollutant. We instrument for PM2.5 utilizing thermal inversion strength.^11^ As revealed in Table 4, the coefficients on PM2.5 remain significant and the magnitudes barely change. The WTPs for a 10 μg/m^3^ reduction in PM2.5 are within a reasonable range, between CNY 52.26 and CNY 59.21 per year.

**Table 4:**
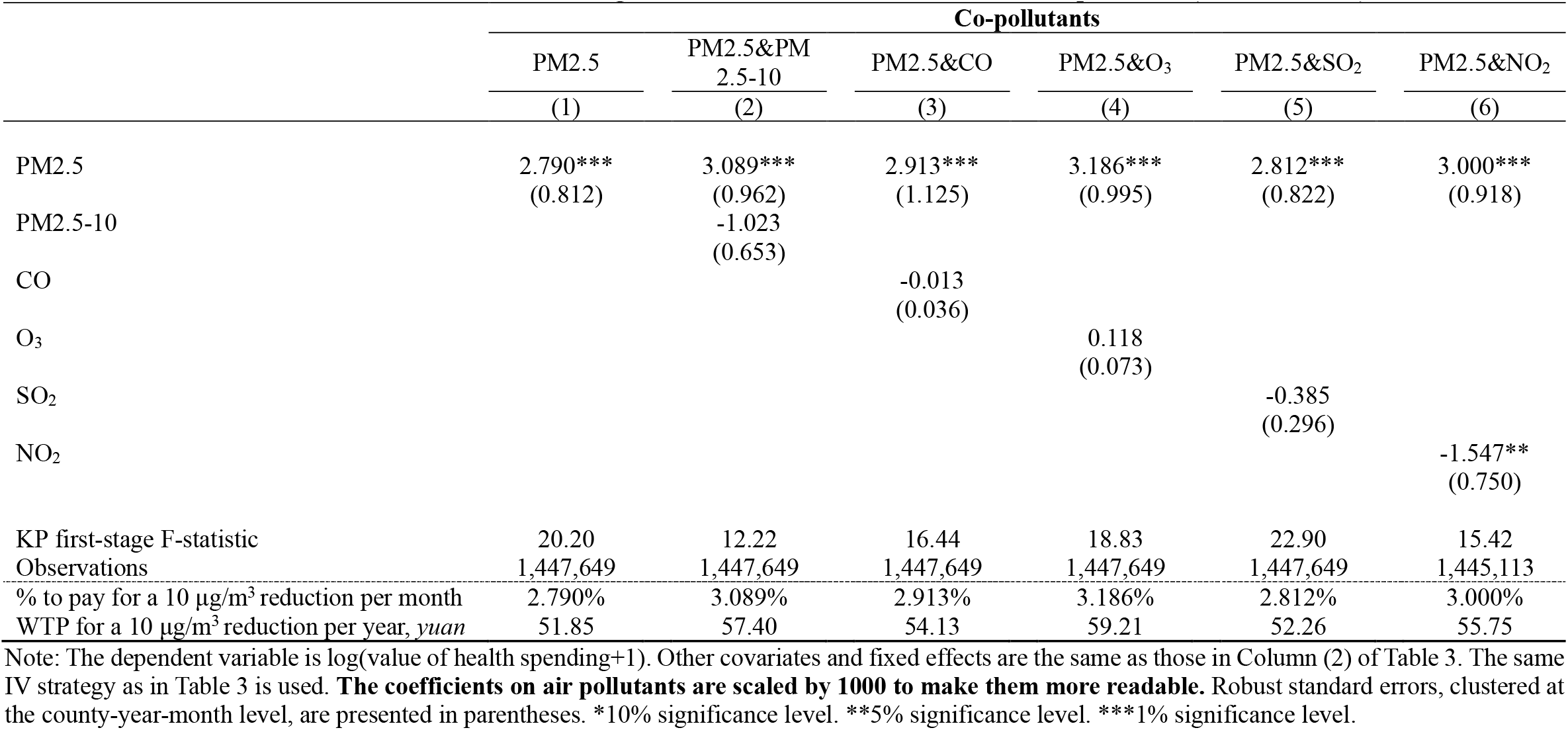
Robustness checks - addressing correlations between PM2.5 and other co-pollutants (2SLS estimates)

|  | Co-pollutants |  |  |  |  |  |
| --- | --- | --- | --- | --- | --- | --- |
|  | PM2.5 | PM2.5&PM<br>2.5-10 | PM2.5&CO | PM2.5&O<sub>3</sub> | PM2.5&SO<sub>2</sub> | PM2.5&NO<sub>2</sub> |
|  | (1) | (2) | (3) | (4) | (5) | (6) |
| PM2.5 | 2.790***<br>(0.812) | 3.089***<br>(0.962) | 2.913***<br>(1.125) | 3.186***<br>(0.995) | 2.812***<br>(0.822) | 3.000***<br>(0.918) |
| PM2.5-10 |  | -1.023<br>(0.653) |  |  |  |  |
| CO |  |  | -0.013<br>(0.036) |  |  |  |
| O<sub>3</sub> |  |  |  | 0.118<br>(0.073) |  |  |
| SO<sub>2</sub> |  |  |  |  | -0.385<br>(0.296) |  |
| NO<sub>2</sub> |  |  |  |  |  | -1.547**<br>(0.750) |
| KP first-stage F-statistic | 20.20 | 12.22 | 16.44 | 18.83 | 22.90 | 15.42 |
| Observations | 1,447,649 | 1,447,649 | 1,447,649 | 1,447,649 | 1,447,649 | 1,445,113 |
| % to pay for a 10 µg/m<sup>3</sup> reduction per month | 2.790% | 3.089% | 2.913% | 3.186% | 2.812% | 3.000% |
| WTP for a 10 µg/m<sup>3</sup> reduction per year, *yuan* | 51.85 | 57.40 | 54.13 | 59.21 | 52.26 | 55.75 |

Note: The dependent variable is log(value of health spending+1). Other covariates and fixed effects are the same as those in Column (2) of Table 3. The same IV strategy as in Table 3 is used. **The coefficients on air pollutants are scaled by 1000 to make them more readable.** Robust standard errors, clustered at the county-year-month level, are presented in parentheses. \*10% significance level. \*\*5% significance level. \*\*\*1% significance level.

Table 5 presents alternative specifications. Column (1) of Table 5 replicates the baseline results in Column (4) of Table 3 for ease of comparison. In Column (2), we perform placebo tests by examining whether “PM2.5 the same month next year” affects health spending. “PM2.5 the same month next year” is instrumented by “thermal inversion the same month next year”. As expected, this variable is not statistically significant.

**Table 5:** Robustness checks - other specifications.

|  | Baseline | Placebo test | Non-linearity | CRE tobit |
| --- | --- | --- | --- | --- |
|  |  | PM2.5 the same month next year |  |  |
|  | (1) | (2) | (3) | (4) |
| PM2.5 | 2.790***<br>(0.812) |  |  | 1.477***<br>(0.312) |
| PM2.5 the same month next year |  | -0.217<br>(0.547) |  |  |
| Non-linearity |  |  |  |  |
| Indicator (PM2.5 ≤ 70 µg/m<sup>3</sup>)<br>(reference group) |  |  | -- |  |
| Indicator (70 < PM2.5 ≤ 100 µg/m<sup>3</sup>) |  |  | 0.030<br>(0.054) |  |
| Indicator (PM2.5 > 100 µg/m<sup>3</sup>) |  |  | 0.174***<br>(0.043) |  |
| KP first-stage F-statistic | 20.20 | 119.1 | 15.34 | -- |
| Observations | 1,447,649 | 1,443,395 | 1,447,649 | 1,447,649 |

Note: The dependent variable is log(value of health spending+1). Other covariates and fixed effects are the same as those in Column (4) of Table 3. In Column (2), the “PM2.5 the same month next year” is instrumented by the “thermal inversion the same month next year”. In Column (3), We use two IVs: thermal inversion strength and the probability of occurrence. **The coefficients on PM2.5 and PM2.5 the same month next year are scaled by 1000 to make them more readable.** Robust standard errors, clustered at the county-year-month level, are presented in parentheses. \*10% significance level. \*\*5% significance level. \*\*\*1% significance level.

Furthermore, we estimate a non-linear relationship between PM2.5 and health spending in Column (3) of Table 5. We classify PM2.5 concentrations into three categories, i.e., PM2.5 ≤ 70 μg/m^3^, 70 < PM2.5 ≤ 100 μg/m^3^, and PM2.5 > 100 μg/m^3^, and assign each category a dummy variable, with PM2.5 ≤ 70 μg/m^3^ being designated as the reference group. We estimate this model utilizing two IVs: thermal inversion strength and the probability of occurrence.^12^ Exposure to heavy air pollution, relative to the reference group, is associated with a significant increase in medical expenditures.

Finally, due to the large number of zeros in the dependent variable outcomes, we check the robustness of our main estimates employing a correlated random effects tobit (CRE tobit) model. As revealed in Column (4) of Table 5, the coefficient on PM2.5 is positive and significant, which suggests that our findings are generally robust when considering the truncated nature of our dependent variable.

### 4.3. Heterogeneous effects

In this section, we examine the heterogeneous effect of air pollution on health spending and estimate the associated WTP across subpopulations. First, we divide the whole sample into six age cohorts of patients (0-5, 6–20, 21–35, 36–50, 51–65, and 66+) and then test the impact of PM2.5 for the six age groups, separately by gender. Table 6 reports the results. Panel A refers to the estimates for males, while Panel B is for those of females. As revealed in Table 6, males are generally more vulnerable to air pollution than their female counterparts. This finding is consistent with the literature, which shows that men’s hedonic happiness and cognitive performance are more affected than women’s (Zhang, Chen, and Zhang 2018; Zhang, Zhang, and Chen 2017a). The young (age 5 years and lower) and the old (age 51 years and above) are more sensitive to air pollution than the middle-aged (21–50 years). Old people (66 years and above) are willing to pay the most for a 10 μg/m^3^ reduction in PM2.5: CNY 374.57 and CNY 212.38 per year for males and females, respectively.

**Table 6:** Heterogeneous effects of PM2.5 on value of health spending, by gender & age (2SLS estimates)

| **A. Male** |  |  |  |  |  |  |
| --- | --- | --- | --- | --- | --- | --- |
|  | 0-5 | 6-20 | 21-35 | 36-50 | 51-65 | 66 and above |
|  | (1) | (2) | (3) | (4) | (5) | (6) |
| *Health spending mean* | 131.69 | 72.10 | 63.40 | 139.81 | 218.62 | 397.43 |
| *SD* | 762.97 | 825.26 | 361.96 | 777.89 | 1147.77 | 2109.55 |
| PM2.5 | 9.193<br>(5.855) | 5.789<br>(3.567) | -0.338<br>(1.524) | 1.723<br>(1.603) | 6.327***<br>(1.946) | 7.854***<br>(2.168) |
| KP first-stage F-statistic | 5.751 | 11.07 | 23.34 | 22.41 | 21.59 | 21.57 |
| Observations | 25,070 | 38,544 | 237,072 | 169,752 | 160,920 | 81,012 |
| % to pay for a 10 µg/m<sup>3</sup> reduction per month | -- | -- | -- | -- | 6.327% | 7.854% |
| WTP for a 10 µg/m<sup>3</sup> reduction per year, *yuan* | -- | -- | -- | -- | 165.99 | 374.57 |
| **B. Female** |  |  |  |  |  |  |
|  | 0-5 | 6-20 | 21-35 | 36-50 | 51-65 | 66 and above |
|  | (1) | (2) | (3) | (4) | (5) | (6) |
| *Health spending mean* | 117.86 | 57.34 | 84.75 | 124.82 | 209.29 | 331.49 |
| *SD* | 945.05 | 604.39 | 485.23 | 622.21 | 944.27 | 2259.13 |
| PM2.5 | 10.293*<br>(6.029) | 4.435<br>(3.360) | -1.814<br>(1.425) | 2.922*<br>(1.568) | 5.062***<br>(1.592) | 5.339**<br>(2.632) |
| KP first-stage F-statistic | 4.705 | 11.42 | 17.25 | 20.38 | 21.47 | 18.95 |
| Observations | 23,763 | 30,744 | 250,728 | 175,188 | 165,228 | 89,628 |
| % to pay for a 10 µg/m<sup>3</sup> reduction per month | 10.293% | -- | -- | 2.922% | 5.062% | 5.339% |
| WTP for a 10 µg/m<sup>3</sup> reduction per year, *yuan* | 145.58 | -- | -- | 43.77 | 127.13 | 212.38 |

Note: The dependent variable is log(value of health spending+1). Other covariates and fixed effects are the same as those in Column (4) of Table 3. The same IV strategy as in Table 3 is used. **The coefficients on PM2.5 are scaled by 1000 to make them more readable.** Each column reports the percentage change in the value of health spending per 10 µg/m<sup>3</sup> increase in PM2.5 concentration. Robust standard errors, clustered at the county-year-month level, are presented in parentheses. \*10% significance level. \*\*5% significance level. \*\*\*1% significance level.

In Panel A of Table 7, we divide health spending into three categories: medication, examination, and treatment. As shown in the panel, health spending in the medication and examination categories is positively correlated with PM2.5. However, only medication expenses are significantly affected by air pollution.

**Table 7:** Heterogeneous effects of PM2.5 on health spending, by spending category, location and health status (2SLS estimates)

|  | A. Spending category |  |  | B. Spending location |  | C. Health status |  |
| --- | --- | --- | --- | --- | --- | --- | --- |
|  | medication | treatment | examination | pharmacy | health facilities | good | poor |
|  | (1) | (2) | (3) | (4) | (5) | (6) | (7) |
| *Health spending mean* | 106.74 | 22.47 | 15.30 | 61.72 | 93.15 | 38.54 | 271.79 |
| *SD* | 519.25 | 387.46 | 183.38 | 291.92 | 984.12 | 123.66 | 1438.13 |
| PM2.5 | 3.215***<br>(0.836) | 0.364<br>(0.364) | -0.126<br>(0.347) | 2.539***<br>(0.772) | 0.847**<br>(0.401) | 2.033**<br>(0.939) | 3.504***<br>(1.059) |
| KP first-stage F-statistic | 20.20 | 20.20 | 20.20 | 20.20 | 20.20 | 20.10 | 19.91 |
| Observations | 1,447,649 | 1,447,649 | 1,447,649 | 1,447,649 | 1,447,649 | 725,714 | 721,935 |
| % to pay for a 10 $\mu\text{g}/\text{m}^3$ reduction per month | 3.215% | -- | -- | 2.539% | 0.847% | 2.033% | 3.504% |
| WTP for a 10 $\mu\text{g}/\text{m}^3$ reduction per year, *yuan* | 41.18 | -- | -- | 18.80 | 9.47 | 9.40 | 114.28 |

Note: The dependent variable is $\log(\text{value of health spending}+1)$ . Other covariates and fixed effects are the same as those in Column (4) of Table 3. The same IV strategy as in Table 3 is used. **The coefficients on PM2.5 are scaled by 1000 to make them more readable.** Each column reports the percentage change in the value of health spending per 10 $\mu\text{g}/\text{m}^3$ increase in PM2.5 concentration. Robust standard errors, clustered at the county-year-month level, are presented in parentheses. \*10% significance level. \*\*5% significance level. \*\*\*1% significance level.

The unique feature of our data is that its health spending records include expenditures in both pharmacies and all levels of health facilities. In Panel B of Table 7, we estimate the effect of PM2.5 on healthcare expenses by spending location, finding that expenditures in pharmacies are much more affected by air pollution. Finding strong responses for medical expenses at pharmacies is particularly important, suggesting that a narrow focus on expenditures in hospitals may result in biased estimates.

Finally, health status may have a significant effect on people’s response to air pollution. Dividing the whole sample into two subsamples by the median value of total medical expenses for each individual, Panel C of Table 7 shows that air pollution has a larger effect on the medical expenditures of people in poor health. The estimates indicate that healthier people are willing to pay 2.033% of their health spending for a 10 μg/m^3^ reduction in monthly average PM2.5, while less healthy people are willing to pay 3.504%. As people in poor health make much larger expenditures, their WTP for a 10 μg/m^3^ reduction in PM2.5 reaches CNY 114.28 per year.

## 5. Discussion

Our preferred specification shows that a 10 μg/m^3^ reduction in monthly average PM2.5 would lead to a 2.79% decrease in the value of health spending and a 0.70% decrease in the number of transactions in pharmacies and health facilities. As the marginal effect of air pollution exposure on total health spending provides a lower bound of people’s WTP for improved air quality, our results indicate that people are willing to pay CNY 51.85 (or USD 8.38) per capita per year for a 10 μg/m^3^ reduction in PM2.5.^13^

To better understand the size of our estimates, in Table A3, we compare our calculated WTP to others in the related literature. Generally, our WTP result is lower than the values estimated using other methods, since our approach provides only a lower bound related to the effect on healthcare spending. For example, Zhang et al. (2017b) find that people, on average, are willing to pay CNY 539 (USD 87.74, or 3.8% of annual household per capita income) per year per person for a 1 μg/m^3^ reduction in PM2.5.

Valuing air quality based on health spending data, Deryugina et al. (2019) find that a 10 μg/m^3^ increase in PM2.5 results in an increase in ER inpatient spending of USD 59.86 per capita per year among the population aged 65 years and older. Williams and Phaneuf (2019) show that a 10 μg/m^3^ increase in PM2.5 results in a 33.1% increase in spending on asthma and chronic obstructive pulmonary disease (COPD). However, all of these studies utilize data from developed countries. Barwick et al. (2018) conducted the first study to examine the morbidity costs of air pollution in China using debit and credit card transactions aggregated at the city level. Their results suggest that the annual household WTP for improved air quality is USD 11.30 for a 10 μg/m^3^ reduction in PM2.5, while ours reveals a larger estimate for WTP, amounting to USD 30.20 per household per year for the same reduction in PM2.5. ^14^ Two issues may contribute to the difference in our WTP estimates. Firstly, our individual-level longitudinal data allow us to remove individual heterogeneity in our estimations, thereby addressing preferences over living environment. Secondly, older persons and low-income residents are more vulnerable to air pollution but less likely to use debit and credit cards, therefore they tend to be excluded from the analysis in Barwick et al. (2018), resulting in potentially underestimated health impact and WTP.

## 6. Conclusion

Previous studies in economics have mainly focused on examining the effects of exposure to air pollution on health factors, such as mortality and hospitalization. Far less is known about the ways in which air pollution affects medical expenditures, especially in developing countries. Our paper is among the first to estimate the morbidity costs of PM2.5 levels by using individual-level health spending data from both pharmacies and health facilities for all age cohorts in China. We employ an IV strategy using thermal inversion as the instrument for PM2.5 in order to address the potential endogeneity in the air pollution measures.

Our analysis shows that PM2.5 has a significant impact on medical expenditures. The estimates suggest that a 10 μg/m^3^ reduction in monthly average PM2.5 leads to a 2.79% decrease in the value of health spending, in addition to a 0.70% decrease in the number of transactions in pharmacies and health facilities. This effect is more salient for males, children (5 years and younger) and older people (age 51 and older) in poor health. Valuing air quality by utilizing health spending data, our estimates suggest that people are willing to pay CNY 51.85 (or USD 8.38) per capita per year for a 10 μg/m^3^ reduction in PM2.5.

In China, the population-weighted annual mean concentration of PM2.5 in 2018 was 51 μg/m^3^, a much larger figure than the annual air quality standards published by the WHO. Our estimates suggest that reducing the annual mean concentration of PM2.5 below that recommended by WHO guidelines (10 μg/m^3^) would enhance welfare by a value equivalent to CNY 212.59 (or USD 32.13) per capita per year.^15^ The optimal environmental regulations depend on the tradeoffs between their costs and benefits. Our valuations of air quality provide useful insights into the benefits of tightening environment regulations.

## Data Availability

Researchers may apply and register for accessing a random representative sample of health spending data via the official webpage of the Wuhan Healthcare Security Administration. http://ybj.wuhan.gov.cn/

## Acknowledgements

Xin Zhang acknowledges financial support from the National Natural Science Foundation of China (72003014). Xun Zhang thanks the National Natural Science Foundation of China (71973014) for financial support. Xi Chen is grateful for financial support from the James Tobin Research Fund at Yale Economics Department, Yale Macmillan Center Faculty Research Award (2017–2019), the U.S. PEPPER Center Scholar Award (P30AG021342, 2016–2018), and NIH/NIA Career Development Award (K01AG053408, 2017–2021). The authors acknowledge helpful comments by participants and discussants at the various conferences, seminars and workshops.

## Appendix A: Supplementary Figures and Tables

**Figure A1:**
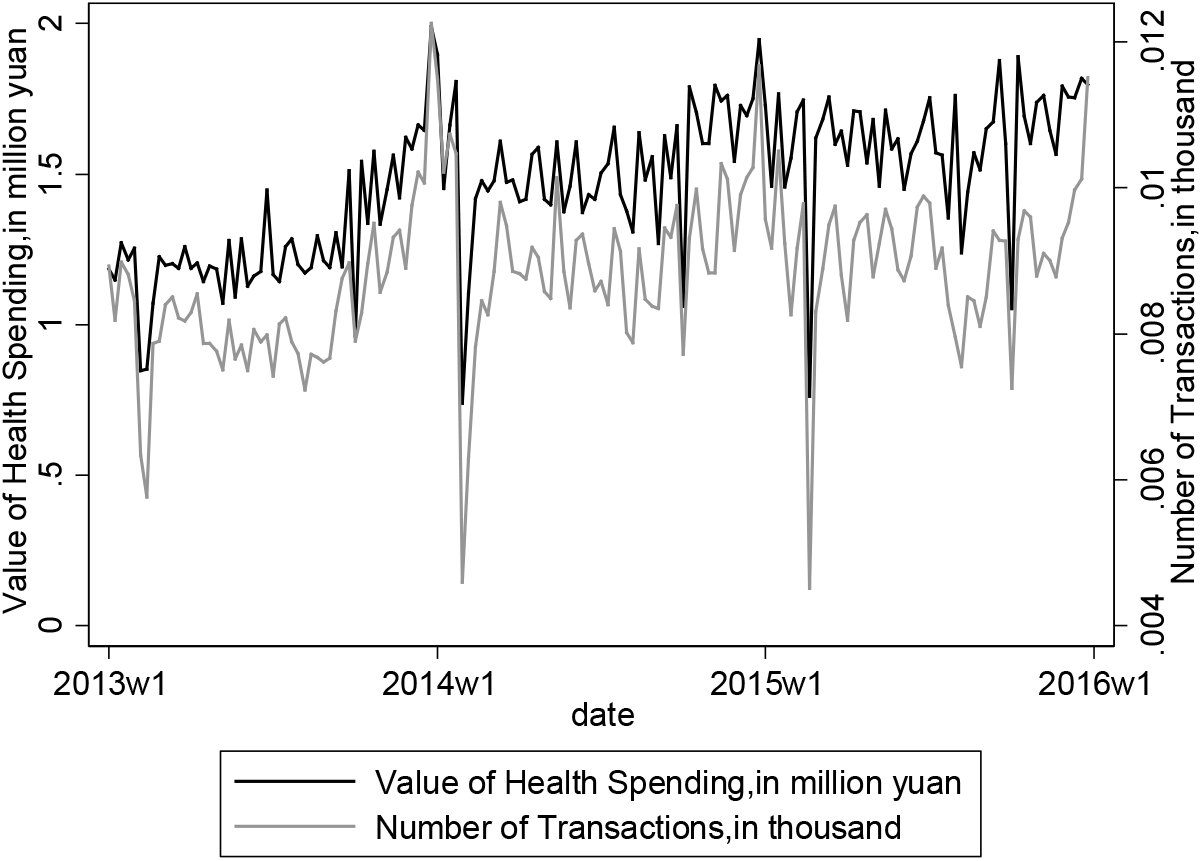
Wuhan weekly health spending, 2013-2015.

**Figure A2:**
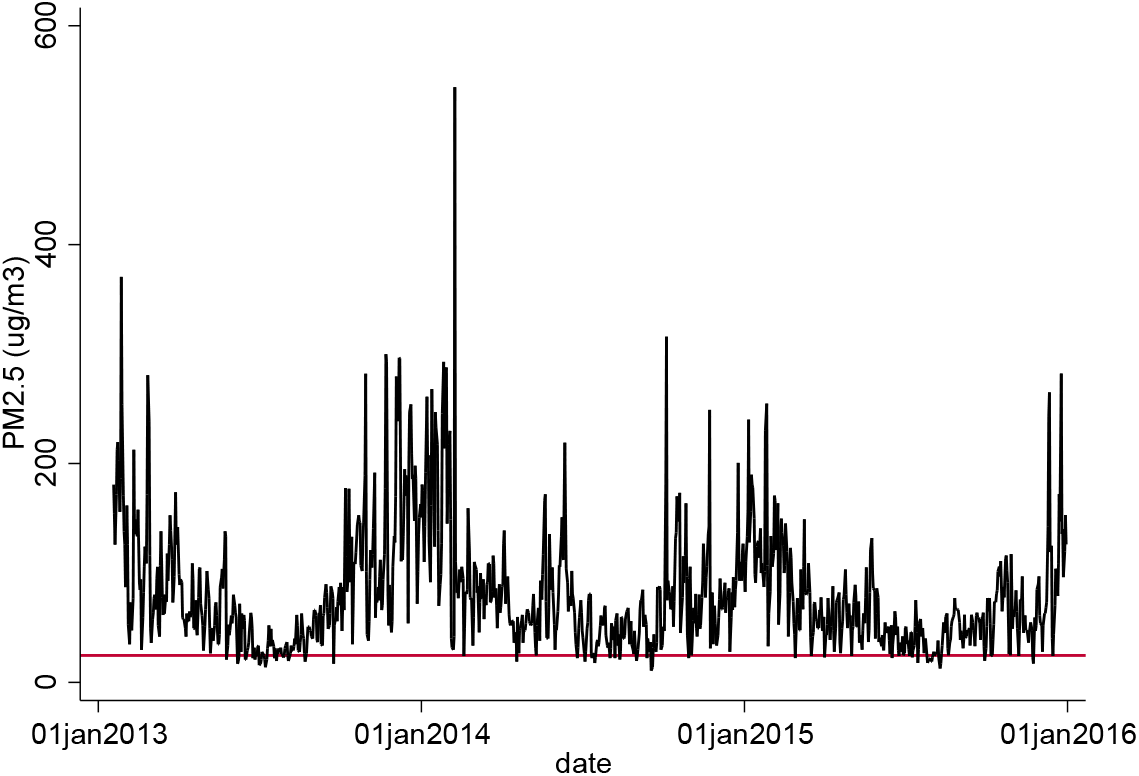
Daily mean PM2.5 (μg/m^3^) in Wuhan city, 2013-2015. Note: PM2.5 = particulate matter with a diameter smaller than 2.5 micrometers. The WHO guidelines level for 24-hour mean PM2.5 is 25 μg/m^3^.

**Table A1:** Correlations between pollutants.

|  | PM2.5 | PM10 | CO | NO<sub>2</sub> | O<sub>3</sub> | SO<sub>2</sub> |
| --- | --- | --- | --- | --- | --- | --- |
| PM2.5 | 1.000 |  |  |  |  |  |
| PM10 | 0.924 | 1.000 |  |  |  |  |
| CO | 0.855 | 0.737 | 1.000 |  |  |  |
| NO<sub>2</sub> | 0.758 | 0.832 | 0.710 | 1.000 |  |  |
| O<sub>3</sub> | -0.188 | -0.165 | -0.216 | -0.225 | 1.000 |  |
| SO<sub>2</sub> | 0.649 | 0.625 | 0.626 | 0.567 | -0.203 | 1.000 |

Note: CO = carbon monoxide. NO<sub>2</sub> = nitrogen dioxide. O<sub>3</sub> = ozone. PM2.5 = particulate matter with a diameter smaller than 2.5 micrometers. SO<sub>2</sub> = sulfur dioxide.

**Table A2:** Effects of thermal inversion on air pollution indicators (first stage)

| Dependent variable | Indicator<br>(70<PM2.5≤ 100 µg/m<sup>3</sup>) | Indicator<br>(PM2.5>100 µg/m<sup>3</sup>) |
| --- | --- | --- |
|  | (1) | (2) |
| **Thermal inversion** |  |  |
| strength, °C | -0.956***<br>(0.175) | 0.642***<br>(0.158) |
| probability of occurrence, % | 0.981***<br>(0.323) | 0.634***<br>(0.230) |
| demographic controls | Yes | Yes |
| weather controls | Yes | Yes |
| individual fixed effects | Yes | Yes |
| year and month fixed effects | Yes | Yes |
| KP first-stage F-statistic | 15.34 | 15.34 |
| Observations | 1,447,649 | 1,447,649 |

Note: The demographic controls include age and its square term. The weather controls include temperature bins (<45 °F, 45-55 °F, 55-65 °F, 65-75 °F, >75 °F), total precipitation, mean wind speed, sunshine duration and relative humidity in polynomial forms. Robust standard errors, clustered at the county-year-month level, are presented in parentheses. \*10% significance level. \*\*5% significance level. \*\*\*1% significance level.

**Table A3:** Summary of WTP from literature.

| **Paper** | **Dose, additional** | **Estimate** |
| --- | --- | --- |
| Deschenes et al. (2007) | 10 percentage points NO<sub>x</sub> | 0.06 percentage point in pharmaceutical expenditures |
| Barwick et al. (2018) | 10 µg/m<sup>3</sup> PM2.5 | 2.6% in hospital visits and pharmacy purchases<br>1.5% in total health expenditures<br>WTP: \$11.3 per household per year |
| Deryugina et al. (2019) | 1 µg/m<sup>3</sup> PM2.5 | \$16.4 thousand per million beneficiaries in ER inpatient spending |
| Williams and Phaneuf (2019) | 1 SD PM2.5 (3.84 µg/m<sup>3</sup>) | 12.7% more spending on asthma and COPD |
| Smith and Huang (1995) | 1 µg/m<sup>3</sup> TSP | WTP: \$98.52 in housing price (in constant 1982-84 dollars) |
| Chay and Greenstone (2005) | 1 µg/m<sup>3</sup> TSP | WTP: \$81-\$213 in housing price (in constant 1982-84 dollars) |
| Bayer et al. (2009) | 1 µg/m<sup>3</sup> PM10 | WTP: \$149-\$185 in housing price (in constant 1982-84 dollars) |
| Ito and Zhang (2016) | 1 µg/m<sup>3</sup> PM10 | WTP: \$1.34 per household per year |
| Welsch (2006) | 1 SD NO<sub>2</sub> (8.238 µg/m<sup>3</sup>)<br>1 SD Lead (0.168 µg/m<sup>3</sup>) | WTP: \$949.50 per capita per year<br>WTP: \$1089.72 per capita per year |
| Levinson (2013) | 1 µg/m<sup>3</sup> PM10 | WTP: \$891 per capita per year |
| Zhang, Zhang and Chen (2017) | 1 µg/m<sup>3</sup> PM2.5 | WTP: 539 CNY per capita per year |
| Wang et al. (2015) | smog mitigation | WTP: 428 CNY per year |
| Sun et al. (2016) | smog mitigation | WTP: 1590 CNY per year |
| Zhang and Mu (2018) | 100-point AQI | 54.5% increase in mask purchases<br>70.6% increase in anti-PM2.5 mask purchases |
| Our estimation | 10 µg/m<sup>3</sup> PM2.5 | 2.79% in the value of health spending<br>0.70% in the number of transactions in pharmacies and health facilities<br>WTP: 51.85 CNY (\$8.38) per capita per year* |

Note: \* 51.85 CNY corresponds to \$8.38 using the average 2013-2015 exchange rate 1 USD = 6.1880 CNY.

## Footnotes

1 There are three main methods of valuing air quality. Each approach has its particular advantages and disadvantages. The hedonic approach infers the value of air quality from property values across regions with differing levels of air pollution exposure (Bayer, Keohane, and Timmins 2009; Chay and Greenstone 2005; Ito and Zhang 2019; Smith and Huang 1995). This approach generally suffers from omitted variable problems, which make the value of air quality endogenous. On the other hand, the contingent valuation method (CVM) directly asks about people’s WTP for better air quality (Sun, Yuan, and Yao 2016; Wang et al. 2015). However, this method is subject to the initial hypothetical monetary value adopted in the survey options and the manner in which questions are framed. The happiness approach calculates the marginal rate of substitution between a reduction in air pollution and household per capita income by holding the happiness constant to assess the monetary value of air pollution (Levinson 2012; Welsch 2006; Zhang et al. 2017b). This approach treats self-reported happiness as a proxy of utility and assumes that utility is comparable among respondents.

2 Their theoretical models illustrate that people’s WTP for clean air can be estimated by adding up different components of the impact of air pollution on population health and behavior. The marginal effect of air pollution on health spending is one of the components, other components include mortality impact, the loss of productivity, and reduction in quality of life.

3 According to the 2018 Environmental Performance Index published by Yale University, the five countries with the most polluted air in the world are Nepal, Bangladesh, India, China, and Pakistan.

4 The UEBMI launched in 1998 as an employment-based insurance program and its coverage reached 92% in 2010. The URBMI launched in 2007 to target the unemployed, children, students, and the disabled in urban areas. It covered 93% of the target population as of 2010 (Yu 2015).

5 As medical expenses covered by the Chinese public health insurance programs are directly billed on medical cards, all the medical payments from people enrolled in public insurance programs – UEBMI and URBMI - are included in the official database by design.

6 The six air pollutant measures are particulate matter with a diameter smaller than 2.5 µm (PM2.5, fine particulates); particulate matter with a diameter smaller than 10 µm (PM10, coarse particulates); carbon monoxide (CO); nitrogen dioxide (NO_2_); ozone (O_3_); and sulfur dioxide (SO_2_).

7 Zhang et al. (2017) suggest that people have a much greater WTP for a reduction in PM2.5 than they do for PM10.

8 Wuhan consists of 13 counties.

9 During the sample period, there were 10 monitoring stations in Wuhan city.

10 Table A1 presents the correlations between air pollutants.

11 We have tried to instrument for PM2.5 and another co-pollutant using thermal inversion strength and the probability of occurrence in Columns (2)-(6) of Table 4. However, we could not pass the weak identification test.

12 See Table A2 for the first-stage estimate.

13 Using the average 2013–15 exchange rate of USD 1 = CNY 6.1880 from the Wind Economic Database.

14 The average number of family members is 3.6 in China (China Family Panel Studies 2014). Therefore, our estimate indicates that the annual household WTP for improved air quality is USD 30.20 for a 10 μg/m^3^ reduction in PM2.5.

15 Using the average 2018 exchange rate of USD 1 = CNY 6.6174 from the Wind Economic Database.

## References

Anderson, Michael L. 2019. “As the Wind Blows: The Effects of Long-Term Exposure to Air Pollution on Mortality.” Journal of the European Economic Association.

Arceo, Eva, Rema Hanna, and Paulina Oliva. 2016. “Does the Effect of Pollution on Infant Mortality Differ Between Developing and Developed Countries? Evidence from Mexico City.” Economic Journal 126(591):257–80.

Barreca, Alan I., Matthew Neidell, and Nicholas J. Sanders. 2017. “Long-Run Pollution Exposure and Adult Mortality: Evidence from the Acid Rain Program.” NBER Working Paper.

Barwick, Panle Jia, Shanjun Li, Deyu Rao, and Zahur Nahim Bin. 2018. “The Morbidity Cost of Air Pollution: Evidence from Consumer Spending in China.” NBER Working Paper.

Bayer, Patrick, Nathaniel Keohane, and Christopher Timmins. 2009. “Migration and Hedonic Valuation: The Case of Air Quality.” Journal of Environmental Economics and Management 58(1):1–14.

Chay, Kenneth, Carlos Dobkin, and Michael Greenstone. 2003. “The Clean Air Act of 1970 and Adult Mortality.” Journal of Risk and Uncertainty 27(3):279–300.

Chay, Kenneth Y. and Michael Greenstone. 2003. “The Impact of Air Pollution on Infant Mortality: Evidence from Geographic Variation in Pollution Shocks Induced by a Recession.” Quarterly Journal of Economics 118(3):1121–67.

Chay, Kenneth Y. and Michael Greenstone. 2005. “Does Air Quality Matter? Evidence from the Housing Market.” Journal of Political Economy 113(2):376–424.

Chen, Shuai, Paulina Oliva, and Peng Zhang. 2017. “The Effect of Air Pollution on Migration: Evidence from China.” NBER Working Paper.

Chen, Siyu, Chongshan Guo, and Xinfei Huang. 2018. “Air Pollution, Student Health, and School Absences: Evidence from China.” Journal of Environmental Economics and Management 92:465–97.

Chen, Yuyu, Avraham Ebenstein, Michael Greenstone, and Hongbin Li. 2013. “Evidence on the Impact of Sustained Exposure to Air Pollution on Life Expectancy from China’s Huai River Policy.” Proceedings of the National Academy of Sciences of the United States of America 110(32):12936–41.

Chen, Yuyu, Ginger Zhe Jin, Naresh Kumar, and Guang Shi. 2012. “Gaming in Air Pollution Data Lessons from China.” B.E. Journal of Economic Analysis and Policy 12(3).

Currie, J. and M. Neidell. 2005. “Air Pollution and Infant Health: What Can We Learn from California’s Recent Experience?” The Quarterly Journal of Economics 120(3):1003–30.

Currie, Janet, Matthew Neidell, and Johannes F. Schmieder. 2009. “Air Pollution and Infant Health: Lessons from New Jersey.” Journal of Health Economics 28(3):688–703.

Deryugina, Tatyana, Garth Heutel, Nolan H. Miller, David Molitor, and Julian Reif. 2019. “The Mortality and Medical Costs of Air Pollution: Evidence from Changes in Wind Direction.” American Economic Review 109(12):4178–4219.

Deschênes, Olivier, Michael Greenstone, and Joseph S. Shapiro. 2017. “Defensive Investments and the Demand for Air Quality: Evidence from the NOx Budget Program.” American Economic Review 107(10):2958–89.

Ebenstein, Avraham, Maoyong Fan, Michael Greenstone, Guojun He, and Maigeng Zhou. 2017. “New Evidence on the Impact of Sustained Exposure to Air Pollution on Life Expectancy from China’s Huai River Policy.” Proceedings of the National Academy of Sciences of the United States of America 114(39):10384–89.

Ghanem, Dalia and Junjie Zhang. 2014. “‘Effortless Perfection:’ Do Chinese Cities Manipulate Air Pollution Data?” Journal of Environmental Economics and Management 68(2):203–25.

Greenstone, Michael and Rema Hanna. 2014. “Environmental Regulations, Air and Water Pollution, and Infant Mortality in India.” American Economic Review 104(10):3038–72.

Grossman, Michael. 1972. “On the Concept of Health Capital and the Demand for Health.” Journal of Political Economy 80(2):223–55.

He, Guojun, Maoyong Fan, and Maigeng Zhou. 2016. “The Effect of Air Pollution on Mortality in China: Evidence from the 2008 Beijing Olympic Games.” Journal of Environmental Economics and Management 79:18–39.

Ito, Koichiro and Shuang Zhang. 2019. “Willingness to Pay for Clean Air: Evidence from Air Purifier Markets in China.” Journal of Political Economy.

Knittel, Christopher R., Douglas L. Miller, and Nicholas J. Sanders. 2016. “Caution, Drivers! Children Present: Traffic, Pollution, and Infant Health.” Review of Economics and Statistics 98(2):350–66.

Levinson, Arik. 2012. “Valuing Public Goods Using Happiness Data: The Case of Air Quality.” Journal of Public Economics 96(9–10):869–80.

Liao, Liping, Minzhe Du, and Zhongfei Chen. 2021. “Air Pollution, Health Care Use and Medical Costs: Evidence from China.” Energy Economics.

Liu, Ya Ming and Chon Kit Ao. 2021. “Effect of Air Pollution on Health Care Expenditure: Evidence from Respiratory Diseases.” Health Economics (United Kingdom) 30(4):858–75.

Long, Ying, Jianghao Wang, Kang Wu, and Junjie Zhang. 2018. “Population Exposure to Ambient PM2.5 at the Subdistrict Level in China.” International Journal of Environmental Research and Public Health 15(12).

Luechinger, Simon. 2014. “Air Pollution and Infant Mortality: A Natural Experiment from Power Plant Desulfurization.” Journal of Health Economics 37(1):219–31.

Moretti, Enrico and Matthew Neidell. 2011. “Pollution, Health, and Avoidance Behavior: Evidence from the Ports of Los Angeles.” Journal of Human Resources 46(1):154–75.

Neidell, Matthew. 2009. “Information, Avoidance Behavior, and Health: The Effect of Ozone on Asthma Hospitalizations.” Journal of Human Resources 44(2):450–78.

Pope, C. Arden and Douglas W. Dockery. 2006. “Health Effects of Fine Particulate Air Pollution: Lines That Connect.” Journal of the Air and Waste Management Association 56(6):709–42.

Schlenker, Wolfram and W. Reed Walker. 2016. “Airports, Air Pollution, and Contemporaneous Health.” Review of Economic Studies 83(2):768–809.

Smith, V. Kerry and Ju-Chin Huang. 1995. “Can Markets Value Air Quality? A Meta-Analysis of Hedonic Property Value Models.” Journal of Political Economy 103(1):209–27.

Sun, Chuanwang, Xiang Yuan, and Xin Yao. 2016. “Social Acceptance towards the Air Pollution in China: Evidence from Public’s Willingness to Pay for Smog Mitigation.” Energy Policy 92:313–24.

Sun, Cong, Matthew E. Kahn, and Siqi Zheng. 2017. “Self-Protection Investment Exacerbates Air Pollution Exposure Inequality in Urban China.” Ecological Economics 131:468–74.

Tanaka, Shinsuke. 2015. “Environmental Regulations on Air Pollution in China and Their Impact on Infant Mortality.” Journal of Health Economics 42:90–103.

Wang, Keran, Jinyi Wu, Rui Wang, Yingying Yang, Renjie Chen, Jay E. Maddock, and Yuanan Lu. 2015. “Analysis of Residents’ Willingness to Pay to Reduce Air Pollution to Improve Children’s Health in Community and Hospital Settings in Shanghai, China.” Science of the Total Environment 533:283–89.

Welsch, Heinz. 2006. “Environment and Happiness: Valuation of Air Pollution Using Life Satisfaction Data.” Ecological Economics 58(4):801–13.

Williams, Austin M. and Daniel J. Phaneuf. 2019. “The Morbidity Costs of Air Pollution: Evidence from Spending on Chronic Respiratory Conditions.” Environmental and Resource Economics 74(2):571–603.

Yu, Hao. 2015. “Universal Health Insurance Coverage for 1.3 Billion People: What Accounts for China’s Success?” Health Policy 119(9):1145–52.

Zhang, Junjie and Quan Mu. 2018. “Air Pollution and Defensive Expenditures: Evidence from Particulate-Filtering Facemasks.” Journal of Environmental Economics and Management 92:517–36.

Zhang, Xin, Xi Chen, and Xiaobo Zhang. 2018. “The Impact of Exposure to Air Pollution on Cognitive Performance.” Proceedings of the National Academy of Sciences of the United States of America 115(37):9193–97.

Zhang, Xin, Xiaobo Zhang, and Xi Chen. 2017a. “Happiness in the Air: How Does a Dirty Sky Affect Mental Health and Subjective Well-Being?” Journal of Environmental Economics and Management 85:81–94.

Zhang, Xin, Xiaobo Zhang, and Xi Chen. 2017b. “Valuing Air Quality Using Happiness Data: The Case of China.” Ecological Economics 137:29–36.

